# Trends and Patterns of Varicella Zoster Virus Infection in Kilifi: A Population-based Serosurvey

**DOI:** 10.64898/2026.02.04.26345611

**Authors:** Antipa K. Sigilai, Caroline N. Mburu, Rose Selim, Rose Ombati, Donald Akech, Boniface Karia, James Tuju, Gaby P. Smits, PGM van Gageldonk, Fiona van der Klis, E. Wangeci. Kagucia, J. Anthony G. Scott, I.M.O. Adetifa

## Abstract

There is limited epidemiologic data on varicella zoster virus (VZV) infections from low- and middle-income countries including Kenya. We aimed to describe the seroepidemiology of VZV in Kilifi, Kenya, where varicella vaccine is not included in the national infant immunization program, in order to generate evidence to inform vaccine policy. We conducted a retrospective serosurvey utilizing archived plasma and serum samples from cross-sectional population-based serosurveys conducted within the Kilifi Health and Demographic Surveillance System between 2009 and 2021. We assayed immunoglobulin G (IgG) for VZV using a validated Luminex multiplex immunoassay and applied a seropositivity cutoff of ≥0.26 International Units per millilitre (IU/mL), as determined by the assay developer. We calculated Bayesian-adjusted age-specific seroprevalence and tested differences in seroprevalence between groups using Chi square. We used a multivariable logistic regression model to estimate associations with VZV IgG antibody seropositivity. We fitted an age-dependent catalytic model to estimate the force of infection (FOI) in children aged 0.5-4, 5-9 and 10-14 years. A total of 2639 samples from children aged <15 years and 546 samples from persons aged ≥15 years were tested. The overall population-weighted seroprevalence of VZV IgG antibodies among children aged 0-14 years was 38.4% (95%CI 27.5-49.5). Age-specific seroprevalence rose from 13.3% (95%CI 5.8-21.6) in children aged 0-4 years to 60.9% (95%CI 45.0-76.2) in those aged 10-14 years. Survey year and age were associated with VZV IgG antibody seropositivity. Children aged 5-9 years had the highest FOI (0.098; 95%CI 0.077-0.120) per susceptible year while mean age of infection was 24.3 years (95%CrI 17.6–30.1). Approximately 40% of individuals entering adulthood in Kenya remain susceptible to VZV infection, suggesting a substantial and underappreciated risk of severe VZV disease in older population including pregnant women. An infant varicella immunization program might avert disease across both paediatric and adult populations.

## Introduction

Primary infection with varicella zoster virus (VZV) causes varicella (chicken pox) which is often benign and self-limiting in healthy individuals, however, it can cause severe disease in neonates, adolescents, pregnant women, adults, and immunocompromised individuals (1–3). VZV infection in the first 20 weeks of pregnancy is of particular concern because of the risk of varicella congenital syndrome that is linked to fatal foetal malformations (4). Rarely, varicella may be characterized by skin and soft tissue bacterial infections, pneumonia, otitis media, central nervous system infections, and even death, particularly in immunocompromised individuals (5–8). Annually, varicella is estimated to cause 4.2 million hospitalizations worldwide out of which approximately 0.1% result in death. VZV can also remain dormant for years in the sensory nerve ganglia, causing herpes zoster (shingles) when reactivated, which mainly affects the elderly and immunocompromised individuals. The global incidence of shingles in adults aged ≥70 years is estimated to be 918.2 per 100,000 person years (9).

Existing population-based serological studies on VZV are predominantly from high income countries (HICs) where routine childhood varicella vaccination is common(10). Most of these studies have consistently reported a high seroprevalence of ≥90% by entry to adolescence, primarily driven by childhood vaccination (11–13). The widespread adoption of universal varicella vaccination in national immunization programs in many HICs has been linked to reduced disease burden and hospitalisation. In Europe for instance, it is estimated that there would be 5.5 million varicella cases every year with up to 3.9 million primary care visits if there were no routine varicella immunization programs (14). In contrast, VZV infection and the potential benefits of varicella vaccine have received little attention in many low- and middle-income countries (LMICs) despite evidence linking VZV infection to missed school days among infected children and significant loss of productivity among caregivers as they provide care to their sick children (15).

The few studies on the epidemiology of VZV in sub-Sahara Africa (sSA) are limited to hospitalised patients, immunocompromised individuals, refugee camp residents, health workers, and pregnant women(16–19). In these studies, the seroprevalence of VZV in children below 5 years of age has been reported to be as low as 8% in the Democratic Republic of Congo(20), 24% in hospitalised Kenyan children aged 1-12 years (21), 45% in children aged 1-4 years in Guinea Bissau (22), and 66% in 4–15-year-old children in Nigeria (23). None of these studies has examined the seroprevalence of VZV and its transmission dynamics among healthy individuals across all age groups at the population level. Understanding population-level VZV immunity and its transmission dynamics can quantify past infections, assess potential risks of outbreaks, and guide country-specific policies to control varicella virus disease. The current study aimed to determine age-specific immunity profiles for VZV in healthy children and adults, underlying transmission patterns, and associated risk factors in Kenya.

## Materials and Methods

### Archived samples

We obtained archived plasma and serum samples from three serosurveillance studies that were conducted within the Kilifi Health and Demographic Surveillance System (KHDSS) between 2009 and 2021. The KHDSS is located along the Kenyan coastline and has records of births, pregnancies, migration, and deaths that have been collected routinely since 2000 when it was established (24). The archived samples were collected biennially as part of malaria cross-sectional surveys conducted between 2009 and 2013 (3 total surveys), annually as pneumococcal serosurveys conducted between 2015 and 2019 (3 total surveys), and between December 2020 to May 2021 as part of a COVID-19 serosurvey. Detailed descriptions of the methodologies used in these surveys have been previously published (25–27).

The malaria cross-sectional surveys enrolled samples of about 500 children aged 0-14 years obtained from randomly selected households in mapped clusters for each survey year. Each of the pneumococcal serological surveys enrolled a cross-sectional, age-stratified, simple random sample of 50 children in each of 10 age categories (i.e., 0, 1, 2, 3, 4, 5, 6, 7, 8-9, and 10-14 years) derived from the Kilifi HDSS database. The COVID-19 serosurvey included a cross-sectional, age-stratified, simple random sample of 850 children and adults from the Kilifi HDSS; 100 children were sampled in each five-year age band below 15 years of age, 50 individuals in each five-year age band between 15 and 64 years of age, and 50 adults aged ≥65 years. The archived samples were de-identified and linked to the metadata using unique identifiers. Therefore, all the investigators had no access to information during or after the study that could link individual participants to the samples.

### Laboratory analyses

Samples were tested for VZV immunoglobulin G (IgG) antibody using a fluorescent-bead-based multiplex immunoassay on the Luminex® MAGPIX® System, alongside six other antigen targets: diphtheria, pertussis, tetanus, measles, mumps, and rubella. This immunoassay was developed and validated by the National Institute of Public Health, and the Environment (RIVM), in The Netherlands (28, 29). Samples were retrieved between August 2021 and January 2023 and testing was conducted at the KEMRI-Wellcome Trust Research Programme Laboratories in Kilifi.

The VZV antigen, VZ-10 ZV strain (Genway GWB-23D26F, California U.S.A), was coupled to coloured magnetic beads. The samples, standards, and controls were diluted in Phosphate-Buffered Saline containing 0.1% Tween 20 and 3% Bovine Serum Albumin. The samples were then diluted in duplicates at ratios of 1:200 and 1:4000. Each plate included in-house controls, blanks, and standards. We used the international rubella standard serum RUBI-1-94 (NIBSC, Potters Bar, United Kingdom) as a reference, which contains a VZV-specific IgG concentration of 22 international units per millilitre (IU/mL). Antibody concentrations were determined by interpolation into a 5-parameter fit standard curve using Milliplex Analyst version 3.55. Antibody levels were expressed in IU/mL; a seropositivity threshold of ≥0.26 IU/mL as determined by RIVM was applied (30).

### Statistical analyses

We first calculated the overall crude seroprevalence stratified by survey type, age, and sex. To enable comparison of seroprevalence across survey years while accounting for age distribution differences, we standardised our estimates by applying multilevel regression and post-stratification(MLRP) adjusted for specificity of the assay. We used the average population structure from the entire series as our standard population for each of the yearly samples to standardize both the age-specific seroprevalence values as well as the total KHDSS population values (**Table S2**). The sensitivity of the assay was given as 100% and specificity was 91.3%. The MLRP was implemented via a Bayesian logistic regression that included age as a covariate. Non-informative priors were used for all parameters, and the model was fitted using rjags (31). This standardisation yielded age and test-adjusted estimates of VZV IgG seroprevalence for each survey year along with associated 95% confidence intervals. Geometric Mean Concentration (GMCs) of VZV IgG antibodies per year and across all ages and their corresponding 95%CI were also calculated and visualised using boxplots.

The samples were grouped by age as follows: 0-4, 5-9, 10-14, 15-24, 25-34, 35-44, 45-54, 55-64, and ≥65 years. Differences in seroprevalence across age groups, sex, and survey year were assessed using the Chi-square test. Independent t-test and one-way analysis of variance (ANOVA) tests were used to determine if there were any significant differences in participants’ mean age by sex, survey years, and type of survey. A multivariate logistic regression model with age, sex, and survey years as independent variables was used to assess risk factors associated with VZV IgG antibody seropositivity.

### Determining age-specific force of infection, basic reproduction number(**R_0_**) and herd immunity threshold

We used a simple catalytic model to estimate the age-specific force of infection (FOI), λ(a), a measure of the incidence of infection in a susceptible population for three age groups: 0.5-4, 5-9, and 10-14 years between 2009 and 2021. Infants who were 6 months or younger were assumed to be protected by maternal antibodies in line with previous FOI studies for varicella (32). To avoid this confounding factor, data for children aged ≤6 months were excluded from the FOI analysis. The model shown below follows individuals from birth and assumes a constant FOI over time independent of age.

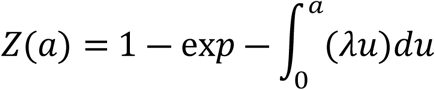

We extended the model to allow for age-varying FOI, and the equations below were modelled:

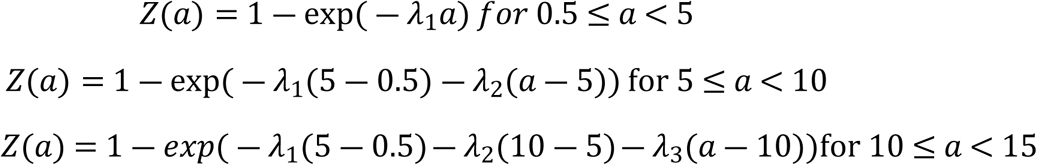

Where *Z*(*a*) is the proportion seropositive at age a while λ_1_, λ_2_, λ_3_ are the age-dependent FOIs. To estimate transmission intensity across the full age range, we extended the model to adults using the 2021 serosurvey by stratifying data into the following age bands: 15–24, 25–34, 35–44, 45–54, 55–64, and ≥65 years. In this extended formulation, the force of infection was assumed piecewise constant within each age interval, such that;

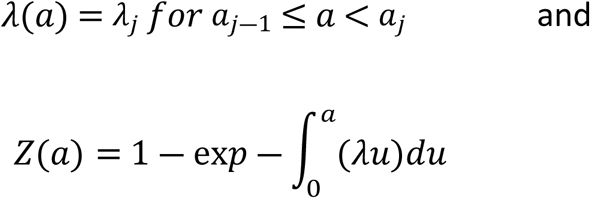

We assumed a closed population, lifelong immunity after varicella infection, and that mortality for infected individuals was similar to that of susceptible individuals. The catalytic model was fit to seroprevalence data using Bayesian methods. Markov chain Monte Carlo (MCMC) and Gibbs sampling algorithm were used to estimate the age-specific FOI assuming a binomial likelihood and non-informative priors between 0 and 1 for the parameters in rjags software. The Gelman-Rubin statistic was used to evaluate convergence.

We estimated the basic reproduction number (*R*_0_) using the standard endemic relationship between the mean age at infection and transmissibility (33). Specifically, we combined age-specific forces of infection estimated from the pooled childhood catalytic model (0–14 years, 2009–2021) with those estimated from the adult catalytic model fitted to the 2021 serosurvey (≥15 years) to reconstruct a piecewise constant force of infection profile over the full age range. The implied mean age at infection (*A*) was computed as;

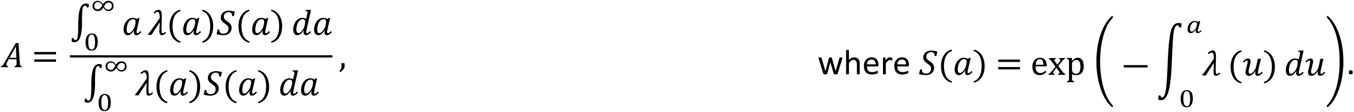

The basic reproduction number was approximated as *R*_0_ = *L*/*A*, where *L* is the average life expectancy at birth over the study period, 61.85 years. Uncertainty in *R*_0_was propagated by Monte Carlo sampling from the posterior distributions of the age-specific forces of infection. From this estimate, we were also able to calculate the herd immunity threshold (HIT) for varicella which is the proportion that would need to be immunised to eradicate endemic transmission of the disease under homogeneous transmission. The HIT was calculated as HIT=1−1/R0

As a sensitivity analysis, we fitted an alternative catalytic model formulation to assess the robustness of the estimated force of infection to modelling assumptions. We extended the current model to allow for waning of immunity following natural infection by including an additional waning parameter with a weakly informative prior (uniform, 0–10% per year) informed by limited published estimates from zoster modelling studies (34). The model was fitted using the same Bayesian framework and likelihood specification as the primary analysis.

### Ethical considerations

Ethical approval to conduct this study was obtained from the Scientific and Ethics Review Unit (SERU) of the Kenya Medical Research Institute (SERU 3847). The surveys contributing the samples tested as part of this study also received approvals from SERU (SSC 1433, SERU 4085, and SSC 1131). All participants in these serosurveys gave individual written informed consent for residual samples to be stored and be used in future research.

## Results

### i. Sociodemographic characteristics of the participants

A total of 2639 samples from children aged <15 years and 546 samples from individuals aged ≥15 years were available for analysis. The number of samples obtained from the malaria cross-sectional surveys were 1072 (33.7%), 1283 (40.3%) from the pneumococcal serosurveys, and 830 (26.0%) from the COVID-19 serosurvey. There was no significant difference in the mean age by sex for all survey years except for the 2021 COVID-19 serosurvey. A summary description of distribution of participants’ mean age and sex per each type of serosurvey across all survey years is presented in **Table 1**.

**Table 1:**
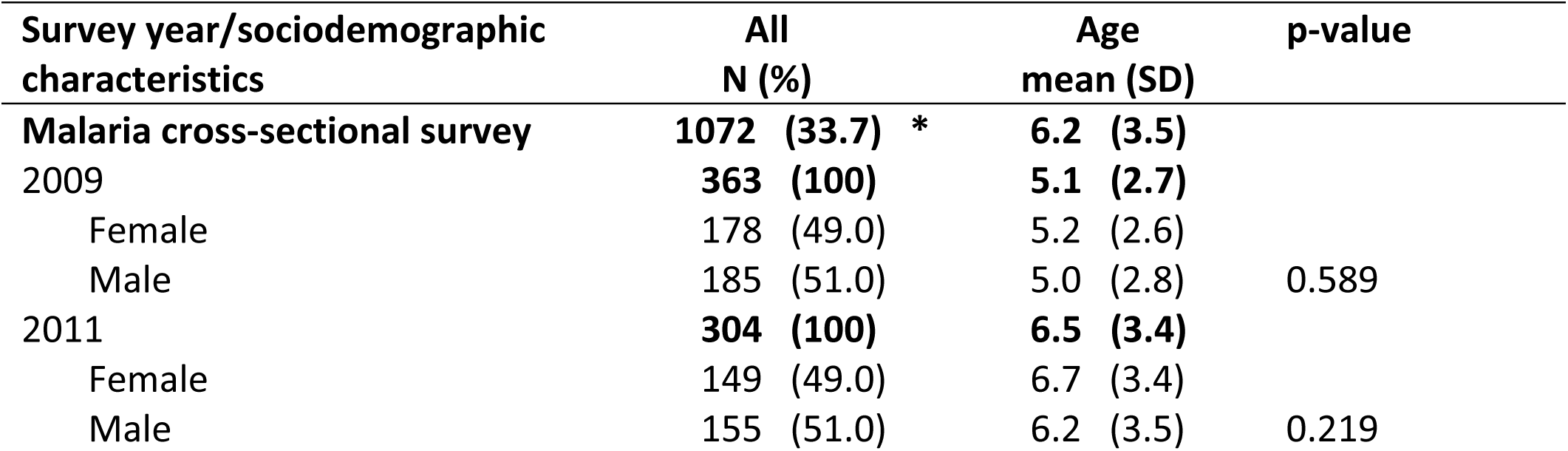
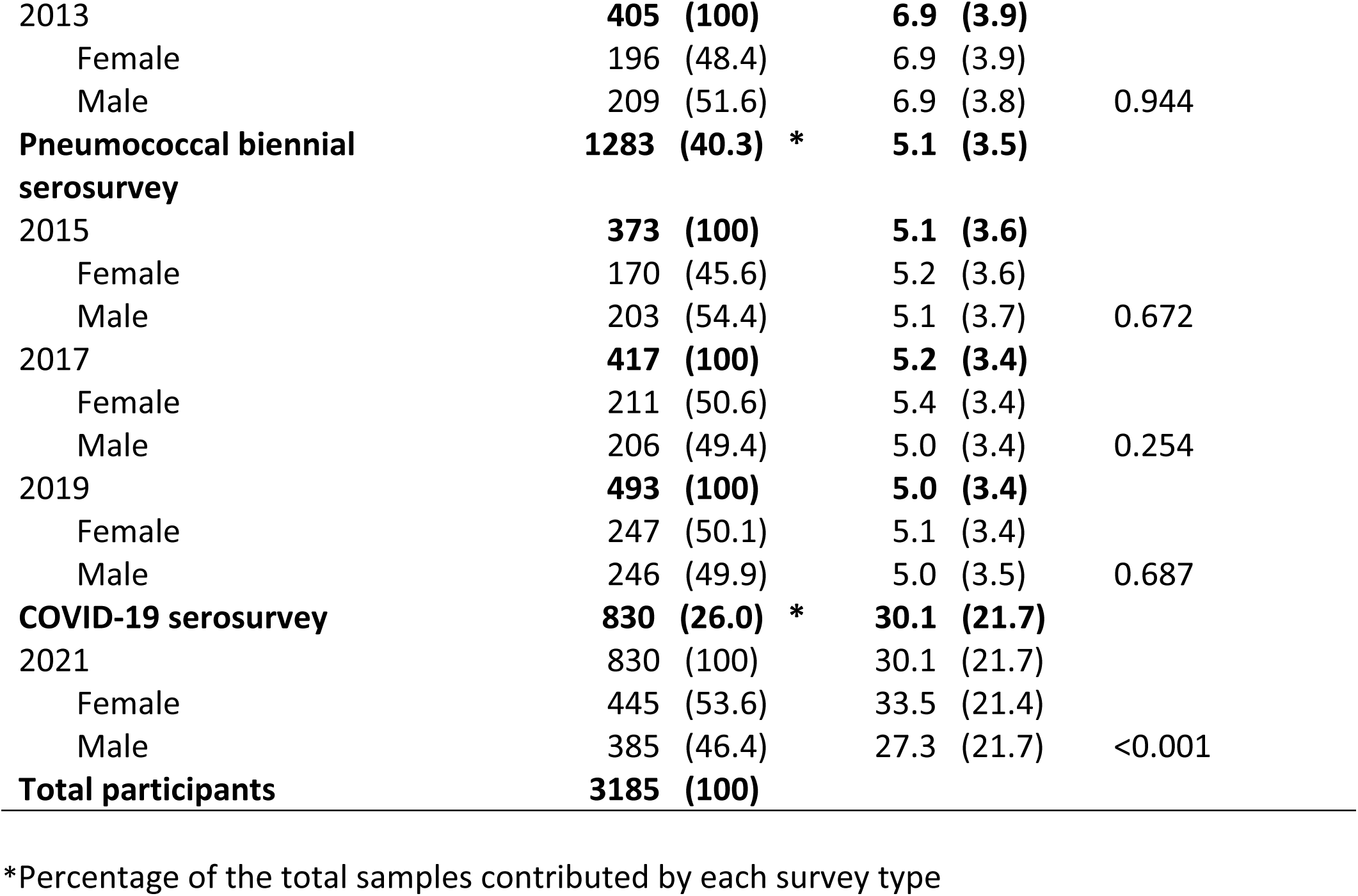
Sociodemographic characteristics and distribution of the participants by each survey type, and sex between. 2009 **and** 2021

| Survey year/sociodemographic characteristics | All N (%) | Age mean (SD) | p-value |
| --- | --- | --- | --- |
| **Malaria cross-sectional survey** | **1072 (33.7) \*** | **6.2 (3.5)** |  |
| 2009 | **363 (100)** | **5.1 (2.7)** |  |
| Female | 178 (49.0) | 5.2 (2.6) |  |
| Male | 185 (51.0) | 5.0 (2.8) | 0.589 |
| 2011 | **304 (100)** | **6.5 (3.4)** |  |
| Female | 149 (49.0) | 6.7 (3.4) |  |
| Male | 155 (51.0) | 6.2 (3.5) | 0.219 |
| 2013 | **405 (100)** | **6.9 (3.9)** |  |
| Female | 196 (48.4) | 6.9 (3.9) |  |
| Male | 209 (51.6) | 6.9 (3.8) | 0.944 |
| **Pneumococcal biennial**<br>**serosurvey** | **1283 (40.3) \*** | **5.1 (3.5)** |  |
| 2015 | **373 (100)** | **5.1 (3.6)** |  |
| Female | 170 (45.6) | 5.2 (3.6) |  |
| Male | 203 (54.4) | 5.1 (3.7) | 0.672 |
| 2017 | **417 (100)** | **5.2 (3.4)** |  |
| Female | 211 (50.6) | 5.4 (3.4) |  |
| Male | 206 (49.4) | 5.0 (3.4) | 0.254 |
| 2019 | **493 (100)** | **5.0 (3.4)** |  |
| Female | 247 (50.1) | 5.1 (3.4) |  |
| Male | 246 (49.9) | 5.0 (3.5) | 0.687 |
| **COVID-19 serosurvey** | **830 (26.0) \*** | **30.1 (21.7)** |  |
| 2021 | 830 (100) | 30.1 (21.7) |  |
| Female | 445 (53.6) | 33.5 (21.4) |  |
| Male | 385 (46.4) | 27.3 (21.7) | <0.001 |
| **Total participants** | **3185 (100)** |  |  |

\*Percentage of the total samples contributed by each survey type

### ii. Age-stratified VZV immunity profiles across survey years

Bayesian age-standardised seroprevalence of VZV IgG antibodies for children aged 0-14 ranged from 28.3% (95% CI 22.0-35.4) in 2017 to 45.6% (95% CI 38.2-53.1) in 2011. The proportion of children considered immune to VZV varied across survey years are shown in **Figure 1A**. A summary of the Bayesian population-weighted and test-adjusted anti-VZV IgG seroprevalence for each survey year is provided in **Table S1**. The geometric mean concentrations (GMCs) of log-transformed VZV IgG antibody showed a cyclic pattern with an initial increase from 0.07 (95%CI 0.06-0.10) IU/mL in 2009 to 0.13 (95%CI 0.10-0.18) IU/mL in 2011. This was followed by a decline to the lowest of 0.04 (95%CI 0.03-0.05) IU/mL in 2017. However, there was a subsequent increase to 0.05 (95%CI 0.04-0.07) IU/mL in 2019 and 0.14 (95%CI 0.11-0.20) IU/mL by 2021 (**Figure 1B**).

**Figure 1:**
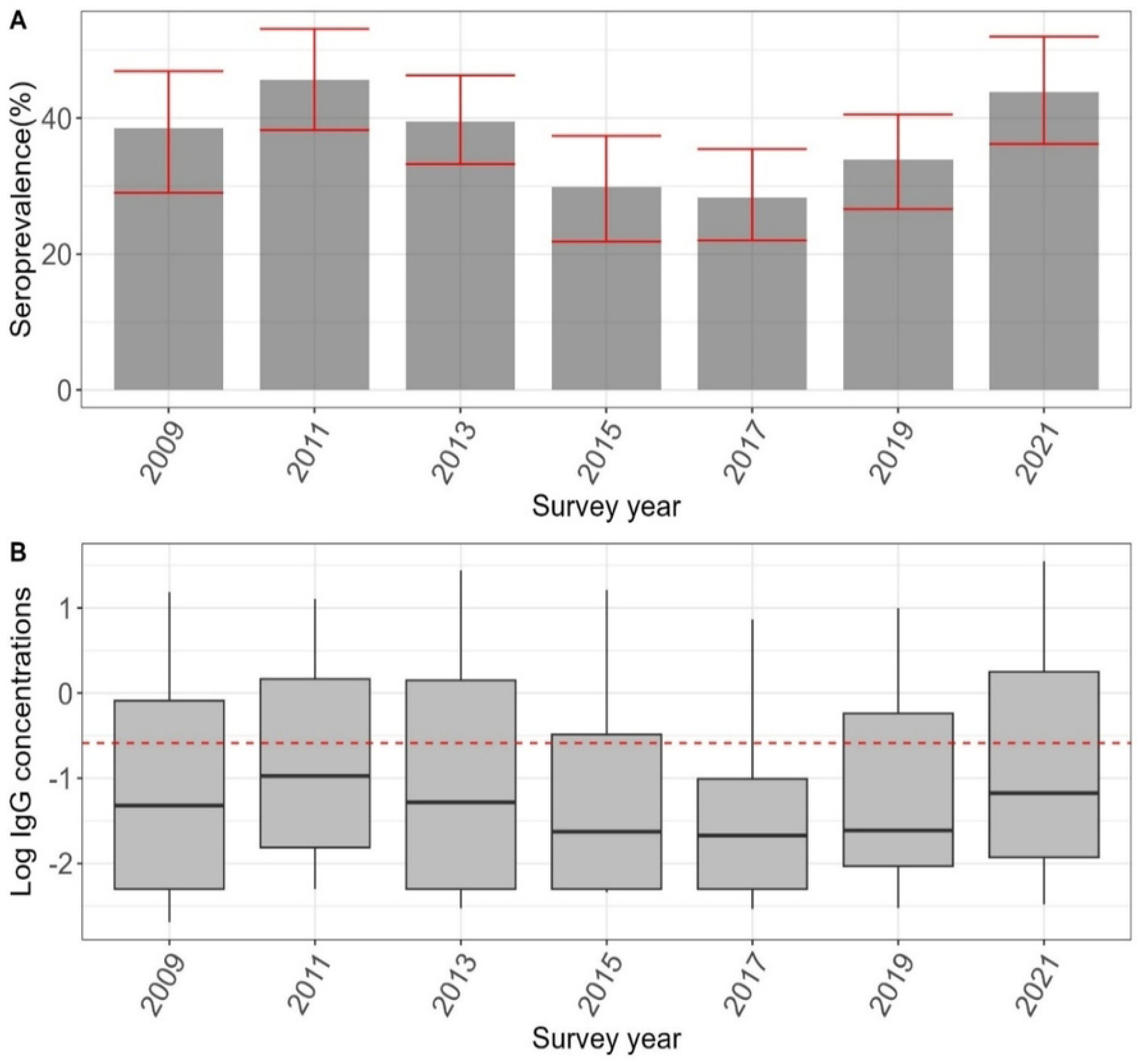
**1A** shows the mean anti-VZV IgG antibody seroprevalence among children aged 0-14 years across each survey year. **1B** shows the log transformed distribution of antibodies with the median indicated by the line in the box and a whisker plot. The red line in figure 1b is the log of the seropositivity threshold for varicella of ≥0.26 IU/mL

Across all the survey years, the Bayesian age-standardised seroprevalence of VZV IgG antibodies was consistently lowest among children aged 0-4 years, ranging from 5.5% (95%CI 1.0-11.4) in 2017 to 25.3 % (95%CI 14.5-36.8) in 2021. In contrast, seroprevalence among children aged 5-9 years ranged from 29.7% (95%CI 20.7-38.6) in 2017 to 55.2% (95%CI 43.1-66.4) in 2021. The highest seroprevalence levels were consistently observed among children aged 10–14-year-olds across survey years with a low of 51.4% (95%CI 34.7-67.4) in 2015 and a peak of 68.3% (95%CI 57.7-79.4; **Figure 2A**) in 2013.

**Figure 2:**
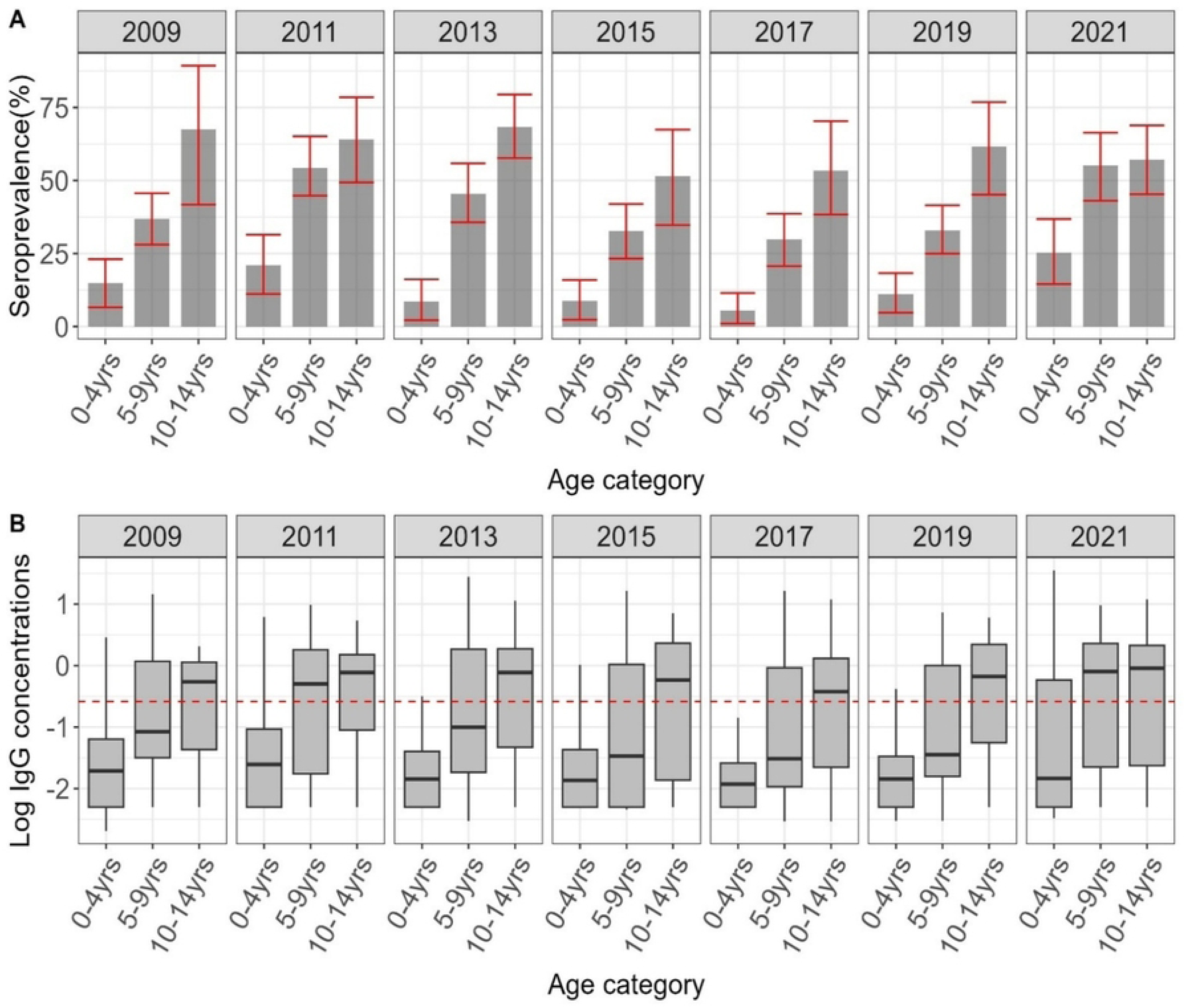
**2A** is the seroprevalence of anti-VZV IgG antibody by age category for children aged 0-14 years for 2009-21 survey years**. 2B** shows the distribution of antibodies with the median indicated by the line in the box and a whisker plot. The red line in the lower figure is the log of the cut-off for seropositivity for varicella of ≥0.26 IU/mL.

Similarly, the GMCs VZV IgG concentration showed an increase with age from 0.03 (95%CI 0.02-0.03) IU/mL among children <5 years to 0.12 (95%CI 0.10-0.13)IU/mL in 5-9-year-olds and reaching 0.28 (95%CI 0.22-0.34) IU/mL in those aged 10-14 years. Overall, the median IgG concentrations of all children <5 years were below seropositivity threshold while those in the 10–14-year group remained consistently above the seropositivity threshold across all survey years. Children aged 5-9 years showed variable findings with the median VZV IgG concentration reaching seropositivity threshold only in 2011 and 2021 (**Figure 2B**). A similar pattern was observed in older children and adults aged ≥15 years in the 2021 serosurvey (**Figure S1A and S1B**).

### Determinants of VZV seropositivity in children

In our multivariate logistic regression model, age was significantly associated with VZV IgG antibody seroprevalence. Compared to children aged 0-4 years, all other age groups had significantly higher odds of testing positive for VZV IgG antibodies (adjusted odds ratios [aORs] 3.71 – 21.92; p< 0.001). Using 2009 as our baseline survey year, the odds of seropositivity were significantly higher in 2011 (aOR= 1.57; 95%CI 1.12-2.19) and 2021 (aOR= 1.48; 95%CI: 1.04-2.09) and lower in 2017 (aOR= 0.65; 95%CI 0.47-0.91; ***Table 2***).

**Table 2:** A multivariate logistic model for risk factors of VZV seropositivity in children aged 0-14 years.

| Independent variables | Participants N | Seropositive n (%) | Unadjusted Odds Ratio (95%CI) | p-value | Adjusted Odds Ratio (95%CI) | p-value |
| --- | --- | --- | --- | --- | --- | --- |
| Age (years) |  |  |  |  |  |  |
| 0-4 | 1059 | 161 (15.1) | 1 |  | 1 |  |
| 5-9 | 1130 | 456 (40.2) | 3.79 (3.09-4.65) | <0.001 | 3.71 (3.02-4.57) | <0.001 |
| 10-14 | 450 | 272 (60.6) | 8.66 (6.73-11.15) | <0.001 | 7.64 (5.90-9.90) | <0.001 |
| 15-24 | 105 (3.3) | 72 (68.6) | 12.28 (7.87-19.16) | <0.001 | 8.06 (4.88-13.33) | <0.001 |
| 25-34 | 100 (3.1) | 71 (71.0) | 13.78 (8.67-21.90) | <0.001 | 9.00 (5.32-15.11) | <0.001 |
| 35-44 | 98 (3.1) | 75 (76.5) | 18.35 (11.17-30.14) | <0.001 | 11.87 (6.83-20.62) | <0.001 |
| 45-54 | 97 (3.0) | 82 (84.5) | 30.76 (17.30-54.70) | <0.001 | 19.96 (10.70-37.24) | <0.001 |
| 55-64 | 98 (3.0) | 84 (85.7) | 33.76 (18.71-60.91) | <0.001 | 21.92 (11.59-41.47) | <0.001 |
| >=65 | 50 (1.6) | 42 (84.0) | 29.54 (13.62-64.09) | <0.001 | 19.26 (8.56-43.32) | <0.001 |
| Survey year |  |  |  |  |  |  |
| 2009 | 363 | 114 (31.4) | 1 |  | 1 |  |
| 2011 | 304 | 141 (46.4) | 1.89 (1.38-2.59) | <0.001 | 1.57 (1.12-2.19) | 0.009 |
| 2013 | 407 | 169 (41.5) | 1.55 (1.15-2.08) | 0.004 | 1.17 (0.85-1.61) | 0.338 |
| 2015 | 373 | 96 (25.7) | 0.76 (0.55-1.04) | 0.089 | 0.76 (0.54-1.07) | 0.114 |
| 2017 | 417 | 100 (24.0) | 0.69 (0.50-0.94) | 0.021 | 0.65 (0.47-0.91) | 0.012 |
| 2019 | 493 | 138 (28.0) | 0.85 (0.63-1.13) | 0.262 | 0.86 (0.63-1.17) | 0.342 |
| 2021 | 284 | 132 (46.5) | 1.90 (1.38-2.61) | <0.001 | 1.48 (1.04-2.09) | 0.028 |

### iii. Force of Infection (FOI)

Both the age-dependent catalytic models in children and adults successfully converged and captured the seroprevalence trends in both children (**figure 3**) and adults (**figure 4**). Among children aged <15 years, the estimated annual rate of infection per susceptible individual was highest among those aged 5-9 years at 0.09 (95%CI 0.07-0.11) followed by children aged 0.5-4 years at a rate of 0.07 (95%CI 0.06-0.08) and lowest in children aged 10-14 years at a rate of 0.04 (95%CI 0.01-0.11). This corresponded to a decrease in reproduction number from 5.75 (95%CI 4.15-6.83) in 5–9-year-olds, to 4.47 (95%CI 4.21-5.36) in under 5-year-olds and further to 3.07 (95%CI 0.25-6.83) in children between 10 and 14 years of age. The FOI in participants who were 15 years or older was lower than 0.05 in all the age groups indicating relatively weak transmission in adulthood (***Table 3***). The implied mean age at infection from the combined pooled childhood (0–14 years) and adult (≥15 years, 2021) force of infection estimates was 24.3 years (95% CrI 17.6–30.1), reflecting delayed acquisition of varicella and substantial residual susceptibility into adulthood. This corresponded to an estimated basic reproduction number of *R*_0_ = 2.54(95% CrI 2.06–3.52). The associated herd immunity threshold was 61% (95% CrI 52%–72%).

**Figure 3:**
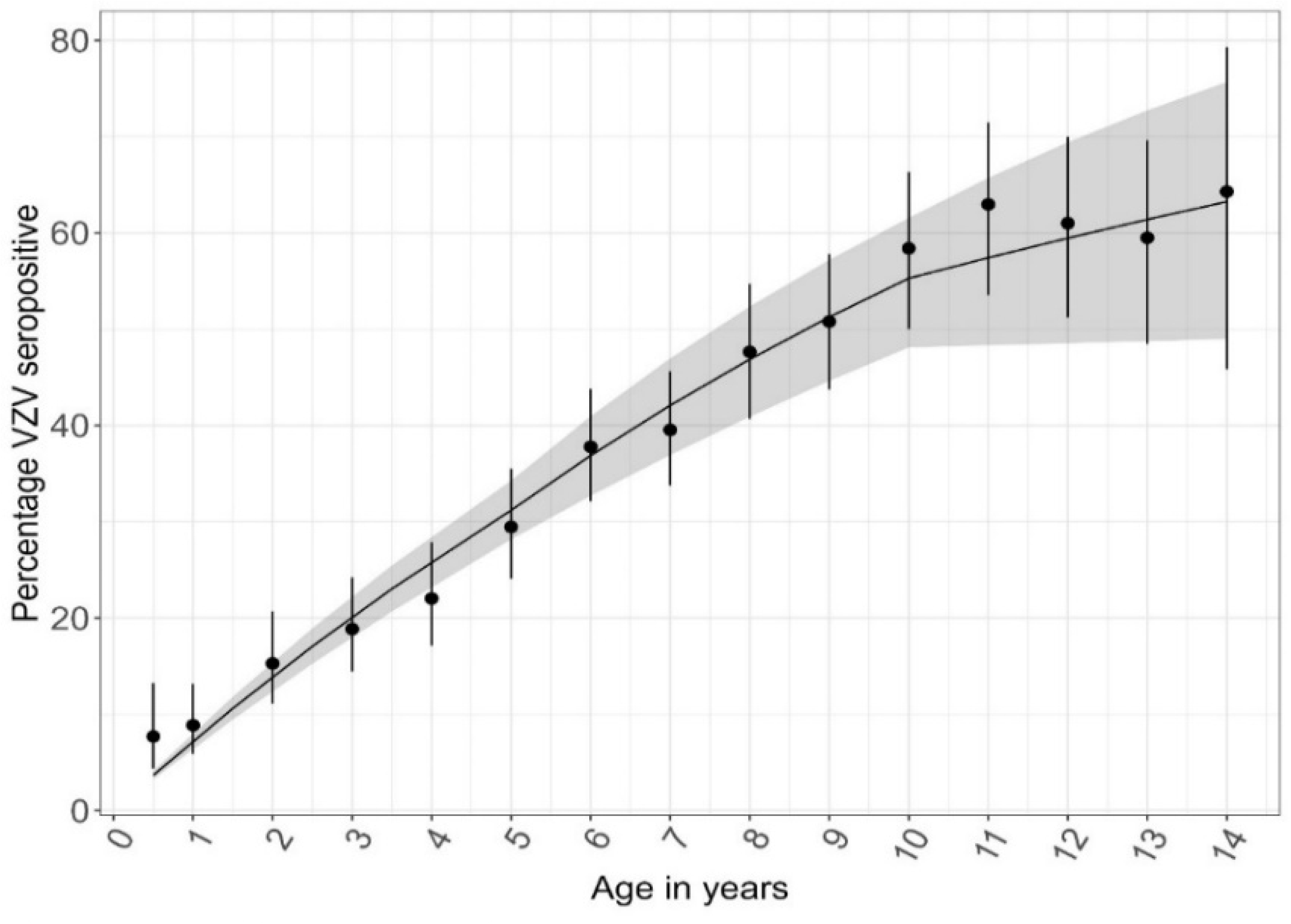
Observed and predicted seroprevalence of IgG antibodies against VZV in Kilifi (2009–2021). Observed point estimates are shown by the dots and the associated binomial 95% confidence intervals. Model predictions are indicated by the solid line.

**Figure 4:**
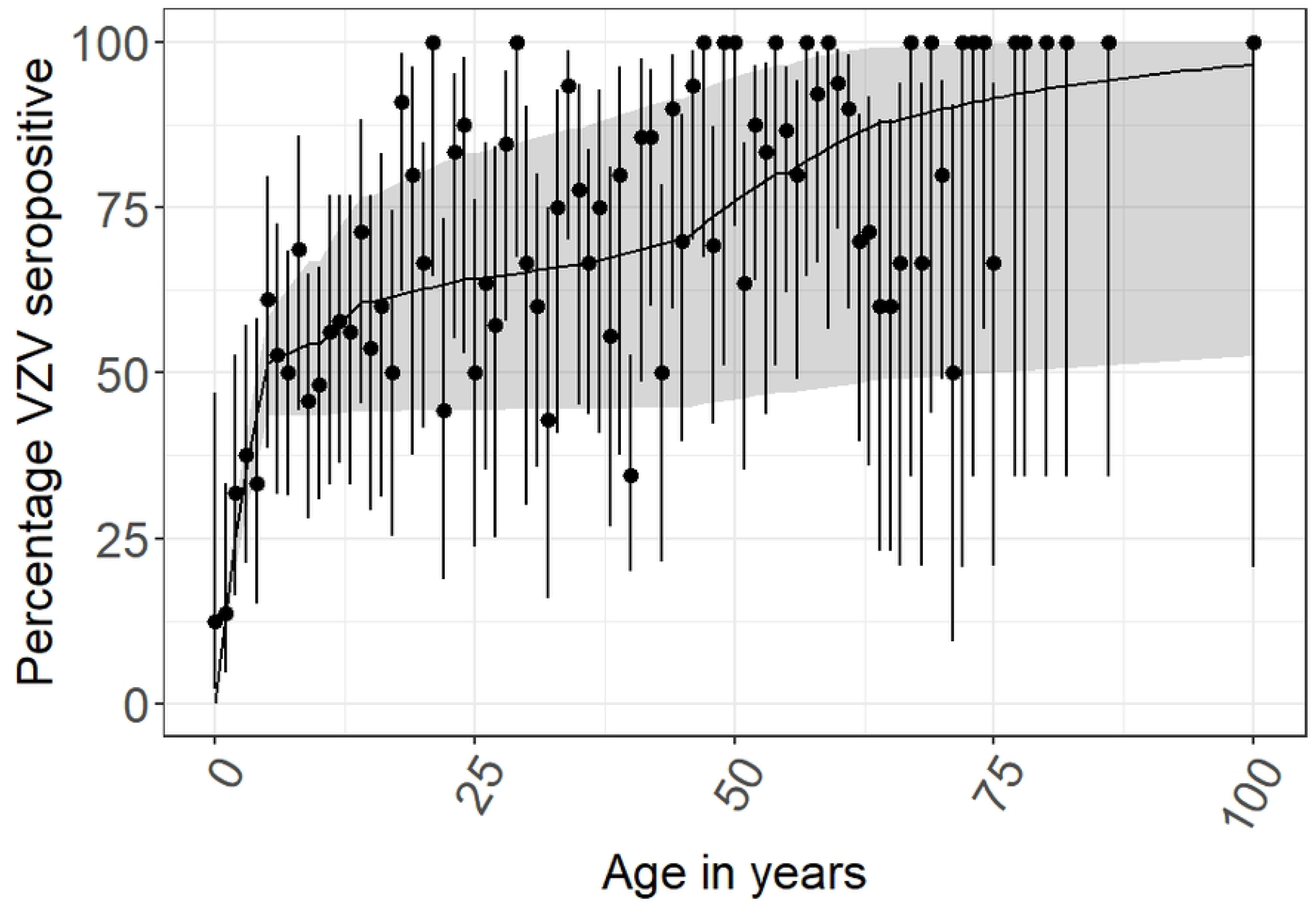
Observed and predicted seroprevalence of IgG antibodies against VZV for 2021 population-wide samples. Observed point estimates are shown by the dots and the associated binomial 95% confidence intervals. Model predictions are indicated by the solid line.

**Table 3:** Force of infection by age estimated from the catalytic model.

| Age Group (Years) | Median | 95%CI |
| --- | --- | --- |
| 0-4 | 0.07 | [0.06-0.08] |
| 5-9 | 0.09 | [0.07-0.11] |
| 10-14 | 0.04 | [0.01-0.11] |
| 15-24 | 0.02 | [0.00-0.05] |
| 25-34 | 0.01 | [0.00-0.05] |
| 35-44 | 0.02 | [0.00-0.07] |
| 45-54 | 0.04 | [0.00-0.07] |
| 55-64 | 0.04 | [0.00-0.15] |
| $\geq 65$ | 0.03 | [0.00-0.11] |

**Note:** The first catalytic model was used for all study years 2009-2021 for children aged 0-14 years while the second catalytic model was applied to 2021 dataset for participants aged >15 years

Inclusion of waning of VZV immunity had minimal impact on the estimated forces of infection across age groups, with FOI estimates remaining highly similar to those obtained under the primary model without waning (**Table S3, Figure S2 and S3**). The posterior median waning rate was 0.087 per year (95% CrI 0.037–0.100); however, the posterior distribution of the waning parameter was broad, indicating that waning was weakly identifiable from these data.

## Discussion

We used representative samples of serological specimens from a biobank to characterize the epidemiology of VZV infection in Kilifi over a span of more than 10 years. Our findings showed that about 38% of children under the age of 15 years had serological evidence of prior VZV infection. VZV IgG antibody seroprevalence increased significantly from about 15% in children aged 0-4 years to 60% among adolescents aged 10-14 years and to >80% among adults aged ≥45years. In contrast, studies in high-income, temperate settings have reported much higher infection-induced VZV seroprevalence. For instance, in the Netherlands infection-induced VZV seroprevalence is about 95% by the age of 6 years (35). This mirrors findings prior to the introduction of an infant varicella immunization program in Germany where VZV seroprevalence was about 86% among children aged 6 years and 94% among those aged 17 years (36).

Still, our findings were comparable with reports from other tropical and sub-tropical countries in the absence of a routine varicella vaccination. For instance, the seroprevalence of VZV IgG antibodies was 50% among Thai children aged 10-14 years and 60% in Singaporean children aged 7-12 years. In Sri Lanka, only 20% of children aged 1-10 years had evidence of VZV infection and 25% of adults aged ≥40 years remained at risk of VZV infection (37). A similar pattern was observed in other tropical and subtropical countries such as Pakistan (38), India (39), and Sudan (40). We also found that the rate of infection was highest in children aged 5-9 years suggesting higher transmission among primary school-going children. This higher transmission observed in children aged 5-9 years in our model is consistent to what has been reported in countries such as Luxembourg (41), Canada (42), Germany, England and Wales among other high-income countries during the pre-vaccine era (43).

Although we observed age-dependent increases in seropositivity due to repeated exposure to naturally circulating VZV, we found that about 40% of persons aged ≥15 years remained susceptible to VZV infection. This pattern is consistent with delayed acquisition of varicella infection and a substantial shift of infections into adolescence and adulthood, as reflected by the relatively high mean age at infection and moderate transmissibility estimated in our models. Although forces of infection in adults were low, a large proportion of individuals escaped infection in childhood and remained susceptible beyond adolescence, resulting in a prolonged tail of infections occurring gradually at older ages.

Available evidence show that delayed exposure to VZV infections in late childhood, adolescence, or early adulthood is associated with more severe disease (44). The sSA region has recorded small to moderate burden of severe VZV illness in adults. Notably, a study from the Democratic Republic of Congo found that of the 124 adults with PCR-confirmed VZV infection, 13-99% had systemic symptoms, 79% had 50 or more lesions, and 15% were bedridden (45). In Zambia, VZV was found to cause about 4% of central nervous system (CNS) disorders among HIV-positive adults, with a 30% case fatality rate among patients with VZV-induced CNS disorders (46). The findings from our study could be incorporated into dynamic VZV transmission models which help build the evidence base for the burden of VZV disease in Kenya.

A significant proportion of adolescents and adults in this study remained susceptible to VZV raising concern of increased risk of severe disease and shingles in older age groups. Although evidence from varicella vaccine effectiveness studies suggest routine childhood varicella immunization programs significantly reduce VZV-associated hospitalisation, complications, and mortality (47–52), local disease burden is needed to justify vaccine introduction in Kenya. An infant immunization program for varicella in Kenya may not only confer direct benefits by averting disease in children but may also contribute towards closing the immunity gap among adults.

As a sensitivity analysis, we explored the potential impact of waning immunity following natural varicella infection. Allowing for waning had minimal effect on the estimated age-specific forces of infection and did not materially alter the overall transmission patterns inferred from the primary analysis. The waning parameter was weakly identifiable from the available cross-sectional serological data, with broad posterior uncertainty, reflecting both limited information in the data and the absence of robust empirical estimates of waning following natural VZV infection in the literature. Although waning could not be reliably quantified in this setting and did not substantially influence inference, loss of immunity following natural infection remains biologically plausible and is thought to occur over long timescales, as evidenced by age-related declines in VZV-specific cellular immunity and reactivation as herpes zoster (34, 53).

### Strengths and Limitations

One of the strengths of this study lies in the random sample of the participants that were drawn from a population with known demographic characteristics. Because of this, we were able to adjust the seroprevalence estimates for the underlying population distribution through population-weighting. This population-based random sampling approach improves the internal validity of the findings. Another key strength of this study is the large sample size that allowed for robust trend analyses of VZV seroprevalence across different ages spanning over 10 years. Therefore, this study provides baseline data on susceptibility patterns which could be used to inform the need for and design of a varicella immunization program in Kenya.

Our study design also had few limitations; first samples for this study were obtained in a rural setting in coastal Kenya limiting the generalization of the findings across Kenya. Second, there were few participants aged <6 months and this limited our understanding of the potential benefits of protection from maternal antibody

## Conclusion

This study describes the seroepidemiology of VZV in children aged 0-14 years and, to a lesser extent, among older children and adults aged ≥15 years in Kenya. Although evidence from this study shows an increase in anti-VZV IgG seropositivity with age, the FOI is low resulting in about 40% of children transitioning into adulthood without VZV immunity. The observed transmission pattern across all ages reflected continued infection into adulthood, suggesting a hidden burden of severe VZV disease among adolescents and adults including sequelae such as congenital varicella syndrome. Taken together, seroepidemiological and modelling data show a significant VZV burden in adults which can be prevented by a childhood vaccine programme. This underscores the need to quantify morbidity burden to guide the evaluation of a varicella vaccine programme in Kenya.

## Data Availability

The raw data files used in this article can be provided upon request through the coordinator of the data governance committee of the KEMRI Wellcome Trust Programme. The committee will review the application and advise as appropriate to ensure that the use of this data is compatible with the consent obtained from participants. Request for the raw dataset should be sent to

## Acknowledgment

We would like to thank all the participants whose samples were used for this work, the Biobank team at the KEMRI-Wellcome Trust Program for assisting in retrieving these samples, and all KWTRP teams involved in the implementation of this work. Special thanks to the KEMIS collaboration for providing additional samples and the RIVM team in the Netherlands for supporting training of our laboratory Research Officer. This paper is published with the permission of the Director, Kenya Medical Research Institute.

## Supplementary Materials

**Figure S1:**
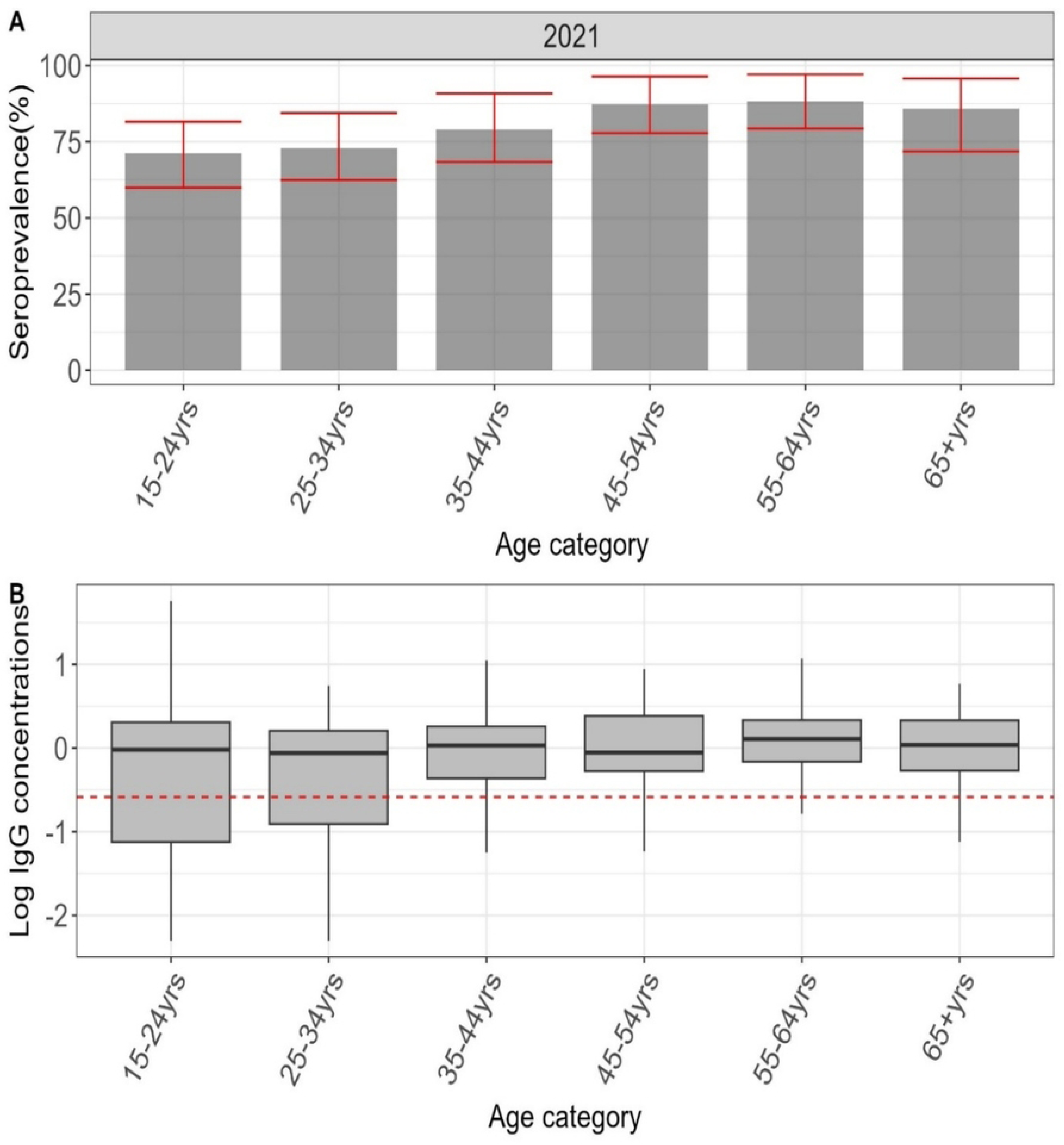
**S1A** Anti-VZV IgG seroprevalence for older children and adults in the 2021 survey. **S1B.** Shows the distribution of VZV GMC IgG levels with the median indicated by whisker plot. The red line in the lower figure is the seropositivity threshold for varicella of ≥0.26 IU/mL.

**Figure S2:**
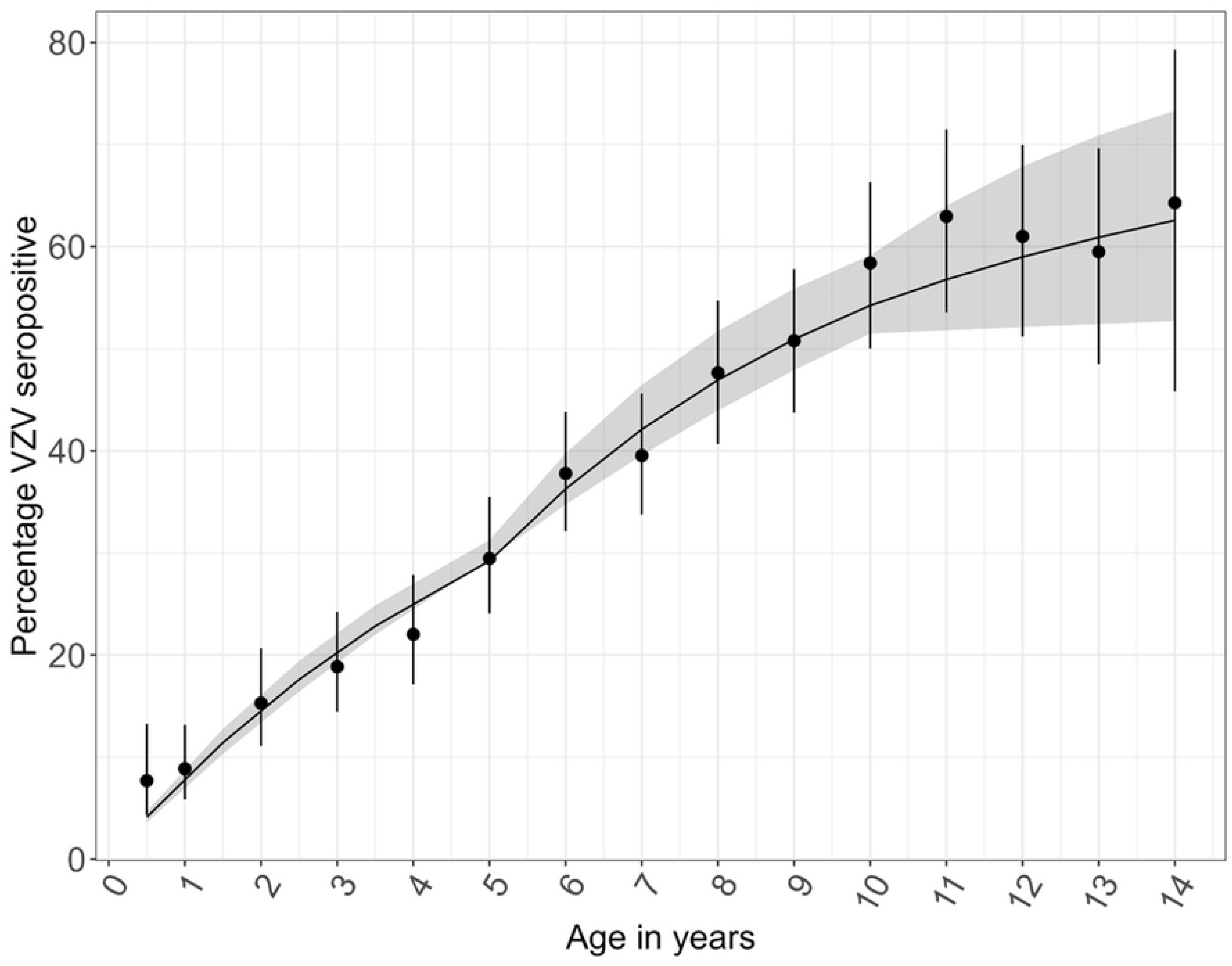
Observed and predicted seroprevalence of IgG antibodies against VZV in Kilifi (2009–2021) from the catalytic model with waning. Observed point estimates are shown by the dots and the associated binomial 95% confidence intervals. Model predictions are indicated by the solid line.

**Figure S3:**
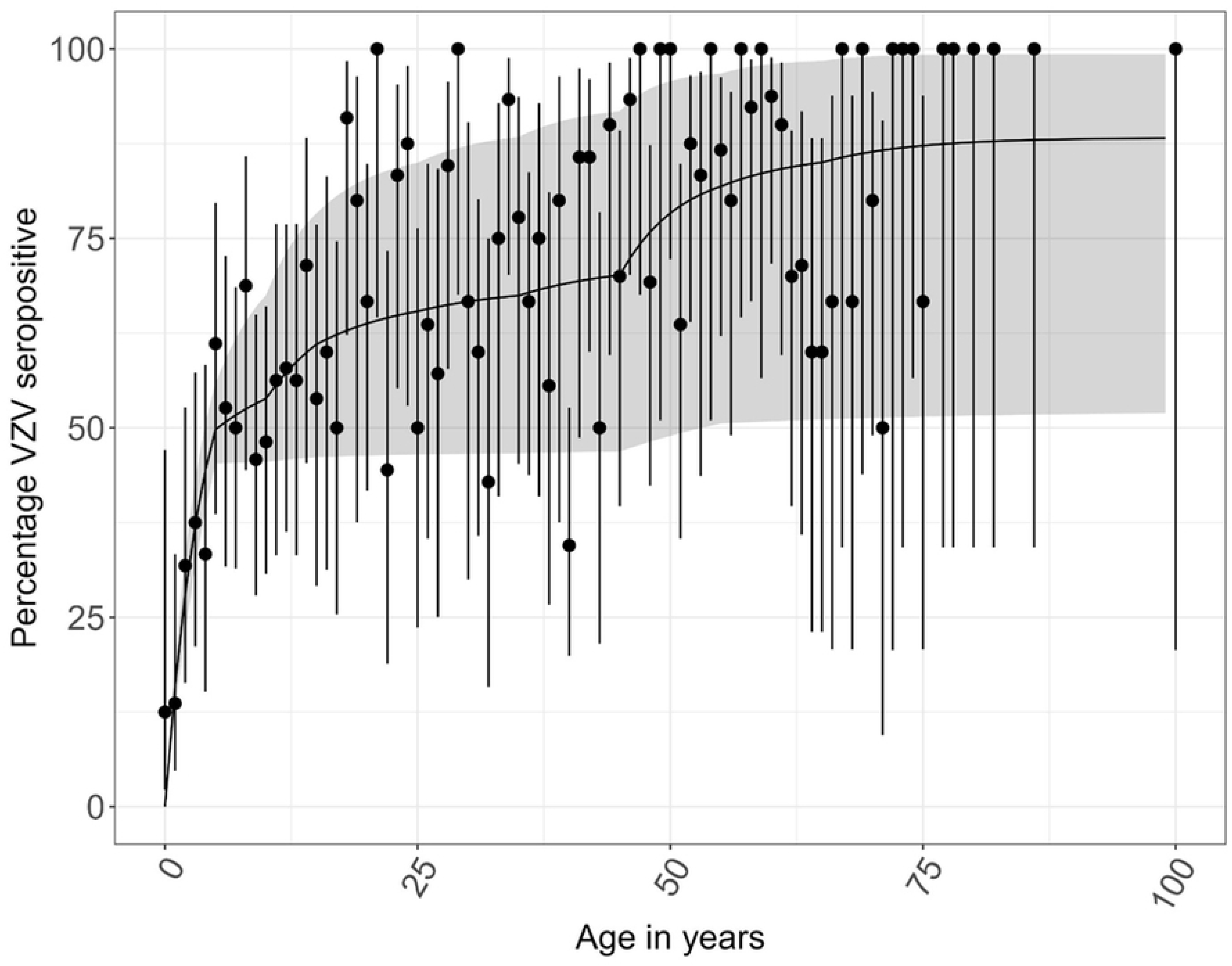
Observed and predicted seroprevalence of IgG antibodies against VZV for 2021 population-wide samples with waning. Observed point estimates are shown by the dots and the associated binomial 95% confidence intervals. Model predictions are indicated by the solid line.

